# Tinnitus and Occupational Noise Exposure among Informal Generator Technicians: Insights from a Nigerian Pilot Study

**DOI:** 10.1101/2025.08.01.25332703

**Authors:** Fatimah Isma’il Tsiga-Ahmed, Nafisatu Bello-Muhammad, Abdulazeez Ahmed

**Author notes:** Corresponding Author: (AA).

## Abstract

**Background:** Tinnitus is a common auditory complaint in chronically noise-exposed individuals and may signal early cochlear dysfunction. In low-resource urban settings where informal work environments dominate, its burden and predictors remain poorly defined. This study explored the prevalence and correlates of tinnitus among generator technicians in Northern Nigeria.

**Methods:** A community-based cross-sectional study was conducted among 73 male generator-repair workers aged 14–57 years. Structured interviews, pure-tone audiometry, and otoacoustic emission (OAE) testing were used to assess sociodemographic factors, noise exposure, and auditory outcomes. Logistic regression identified independent predictors of tinnitus.

**Results:** Prevalence of tinnitus was 45.2%. Significant bivariate associations included marital status (p < 0.001), ≥10 years occupational exposure (p = 0.003), audiometric hearing loss (p = 0.026), and OAE failure (p < 0.001). In multivariable analysis, prolonged occupational exposure (aOR = 4.44) increased odds of tinnitus, while preserved OAE function was protective (aOR = 0.099) for tinnitus.

**Conclusion:** Tinnitus is prevalent among informal generator workers and is strongly linked to prolonged exposure and subclinical cochlear dysfunction. Incorporating OAE screening and hearing health education into community-based programs may enable early detection and prevention of auditory damage in underserved settings.

## INTRODUCTION

Tinnitus, the perception of sound without an external source, and affects an estimated 4.6% to 43% of individuals globally.^[1–6]^ Though its aetiology remains multifactorial, it is widely recognized as a common symptom in populations exposed to prolonged community, leisure or occupational noise.^[7–9]^ Beyond its auditory manifestations, persistent tinnitus can negatively impact sleep, emotional well-being, and overall quality of life, with links to anxiety and depression increasingly being reported.^[10–12]^

In occupational contexts, tinnitus may be an early marker of cochlear dysfunction, often preceding measurable hearing loss.^[13, 14]^ While industrial workers in regulated environments are frequently studied, informal-sector workers, such as generator technicians, welders, and artisans, operate in uncontrolled acoustic conditions, frequently exceeding 90 dB(A) without adequate protection or access to routine hearing screening.^[15–19]^ Despite this heightened vulnerability, the auditory health of informal workers remains severely underexplored, particularly in low-resource urban economies.

In Nigeria, unreliable power supply has spurred widespread reliance on electricity generators, resulting in the proliferation of informal generator-repair businesses concentrated in urban centres.^[18, 20]^ Workers in this setting are regularly exposed to prolonged and intense noise, yet there is limited research on their auditory profiles, especially early symptoms like tinnitus that may reflect subclinical cochlear changes.

Furthermore, emerging literature suggests that tinnitus is shaped not only by noise exposure but also by sociodemographic and psychological factors, including age, education, emotional stress, and beliefs about hearing health.^[21–23]^ Understanding the intersection of these factors in informal noise-exposed populations could inform accessible screening tools and targeted preventive strategies, particularly where conventional diagnostics like audiometry and otoacoustic emissions (OAE) testing are seldom available.

This pilot study aims to assess the prevalence and predictors of tinnitus among informal generator technicians in Northern Nigeria. By examining sociodemographic, occupational, and audiological variables, including pure-tone audiometry and OAE, we seek to identify markers of early auditory dysfunction and highlight opportunities for community-based hearing conservation in underserved work environments.

## MATERIALS AND METHODS

### Study Design and Setting

This was a community-based cross-sectional study conducted in Kano State, Northwestern Nigeria. Data collection spanned a four-month period (4^th^ June – 30^th^ September, 2024), targeting densely populated metropolitan areas characterized by informal electrical generator repair workshops.

### Study Population and Eligibility Criteria

Participants were male generator technicians (a male dominated profession in our clime) registered with the local generator technician’s/mechanics’ union, aged 14–57 years, and actively engaged in repair work at the time of the study. Written informed consent was obtained from all participants. Individuals previously diagnosed with otologic disease or those older than 57 years (to reduce confounding from age-related hearing loss) were excluded.

### Sampling Procedure

A minimum sample size of 60 was calculated using the formula for pilot studies,^[24]^ with an additional 10% buffer for attrition. A two-stage random sampling strategy was applied:

**Stage 1:** Four (4) metropolitan Local Government Areas (LGAs), Nassarawa, Tarauni, Fagge, and Kumbotso, were randomly selected out of a total of eight to enhance reach and feasibility, given that generator technicians in Kano are primarily concentrated in the urban areas.

**Stage 2:** From the union registries obtained in each selected LGA, generator technicians were randomly selected using computer-generated numbers. The number of participants from each LGA was proportionally allocated based on registry size.

### Data Collection Tools and Measures

An interviewer-administered, pretested questionnaire was used to collect data on sociodemographic variables (age, marital status, education, income), occupational exposure (years of experience, daily noise exposure), and tinnitus history. Tinnitus was defined as self-reported perception of ringing, buzzing, or other phantom auditory sensations in one or both ears.

### Hearing Assessment Procedures

Participants underwent basic otoscopic examination followed by two complementary auditory tests:

#### Pure-Tone Audiometry (PTA)

Conducted in a sound-treated booth using a calibrated diagnostic audiometer (Model: Interacoustic AT235H). Audiometric thresholds were measured across 0.5–8 kHz frequencies. Impairment was defined as PTA >25 dB HL.

#### Otoacoustic Emissions (OAE) Testing

Performed using a portable screener (Sentiero-Path Medical) after PTA. Bilateral testing was conducted, and results were categorized as “pass” only when both ears passed. Any unilateral or bilateral failure was considered indicative of cochlear dysfunction.

Ambient noise levels were controlled during audiometric testing to ensure fidelity of threshold detection.

### Rationale for Dual Hearing Assessment

Both PTA and OAE were employed due to their distinct but complementary pathways. While PTA assesses behavioural hearing thresholds, OAEs provide objective insight into outer hair cell function. Several studies have highlighted the enhanced sensitivity and versatility of OAE testing, especially in detecting subclinical cochlear changes that may precede audiometric loss. Their combined use strengthens diagnostic reliability and enables a broader characterization of auditory health.

### Data Analysis

Data were entered into Microsoft Excel and analysed using SPSS version 20 (IBM Corp., Armonk, NY). Frequencies and percentages summarized categorical variables, while continuous data were reported as means ± SD or medians with interquartile ranges depending on distribution (assessed via Kolmogorov-Smirnov test). Chi-square or Fisher’s exact tests assessed bivariate relationships, with independent t-tests or Mann–Whitney U tests for continuous variables. Variables with p < 0.10 were entered into logistic regression to identify independent predictors of tinnitus. Adjusted odds ratios (aOR) with 95% confidence intervals were reported, and a p-value < 0.05 was considered statistically significant.

## RESULTS

A total of 73 generator-exposed workers were ultimately enrolled for this study. Among this cohort, 33 (45.2%) reported tinnitus. The mean age was 34.9 ± 10.2 years. Most participants (64.4%) were aged 26–45 years, 68.5% were married, and 53.4% had at least 10 years of occupational experience. Over 93% reported daily exposure to generator noise exceeding 8 hours (table 1). All the participants were male and reported no use of hearing protection device.

**Table 1.** Bivariate Associations between Tinnitus and Participant Characteristics.

| Variable | Tinnitus Present (%) | Tinnitus Absent (%) | p-value |
| --- | --- | --- | --- |
| Age Group (26–45) | 69.7% | 60.0% | 0.305 |
| Marital Status (Married) | 90.9% | 50.0% | **<0.001** |
| Years in Occupation $\geq 10$ | 72.7% | 37.5% | **0.003** |
| Audiometric Impairment | 63.6% | 37.5% | **0.026** |
| OAE Failure | 54.5% | 12.5% | **<0.001** |
| Family History of Deafness | 15.2% | 12.5% | 0.743 |
| Daily exposure ($>8$ hours) | 100.0% | 87.5% | 0.060 |

Bivariate analysis revealed statistically significant associations between tinnitus and marital status (χ² = 14.023, p < 0.001), years of occupational exposure (χ² = 9.018, p = 0.003), audiometric impairment (χ² = 4.942, p = 0.026), and OAE failure (χ² = 14.813, p < 0.001). Tinnitus was more prevalent among married participants, those with ≥10 years of generator work, and those with abnormal audiometry and OAE outcomes. No significant associations were found with age group, income, education level, eight-hour daily noise exposure or family history of hearing loss (table 1 & figure 1).

**Figure 1.**
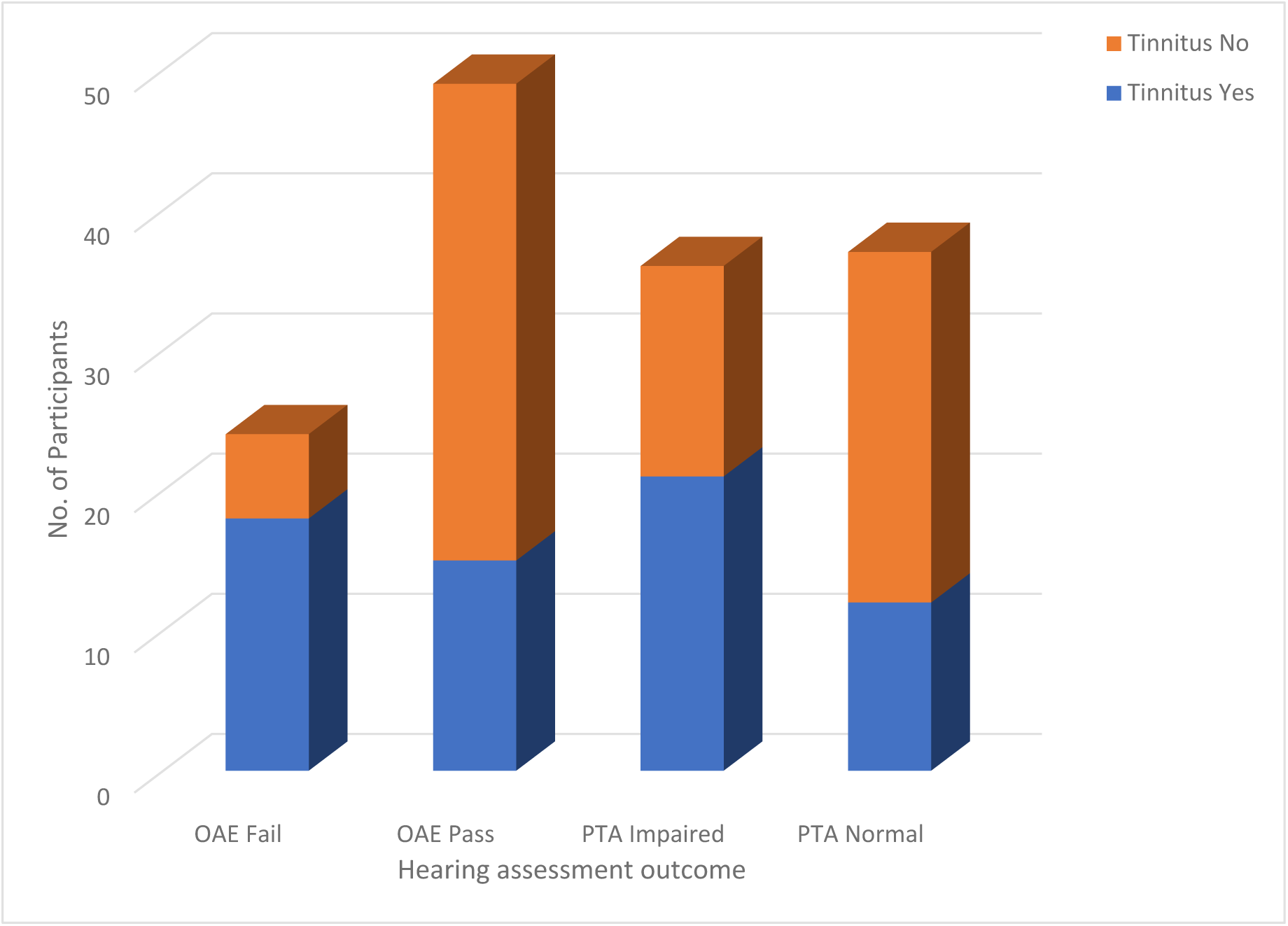
Distribution of Tinnitus Presence by Audiometric and Otoacoustic Emission (OAE) Outcomes among Informal Generator Technicians.

In the logistic regression model, two factors emerged as statistically significant predictors of tinnitus. Technicians with ≥10 years of occupational exposure were over four times more likely to report tinnitus compared to their less-exposed counterparts (aOR = 4.44; 95% CI: 1.04–18.95; p = 0.044). Conversely, participants who passed the OAE screening had substantially lower odds of experiencing tinnitus (aOR = 0.099; 95% CI: 0.020–0.481; p = 0.004) and as such protective, underscoring the protective role of intact cochlear outer hair cell function (table 2). Age, education, audiometric status, daily noise exposure and family history were not significant in the final model. The model explained 33% of the variance (Nagelkerke R² = 0.33), with an overall predictive accuracy of 72.6% and acceptable calibration (Hosmer–Lemeshow p = 0.064).

**Table 2.** Multivariable Logistic Regression for Predictors of Tinnitus.

| Predictor | aOR | 95% CI | p-value |
| --- | --- | --- | --- |
| Years in Occupation $\geq 10$ | 4.44 | 1.04 – 18.95 | 0.044 |
| OAE Failure | 0.099 | 0.020 – 0.481 | 0.004 |
| Other covariates | — | — | >0.05 |

As shown in table 3, tinnitus prevalence varied modestly across age groups, with no clear age-dependent trend. Although age was not a statistically significant predictor in regression analyses, the distribution suggests notable symptom reporting even among younger technicians.”

**Table 3.** Prevalence of Self-Reported Tinnitus by Age Group among Informal Generator Technicians in Kano, Nigeria.

| Age Group (Years) | No. with Tinnitus | Total in Group | Prevalence (%) |
| --- | --- | --- | --- |
| 14–25 | 3 | 12 | 25% |
| 26–35 | 23 | 47 | 48.9% |
| $\geq 45$ | 7 | 14 | 50% |
| Total | 33 | 73 | 45.2% |

A cross-tabulation of PTA and OAE results by tinnitus status is shown in table 4. Of note, all participants with normal PTA but failed OAE responses reported tinnitus, underscoring the diagnostic value of subclinical cochlear testing.

**Table 4.**
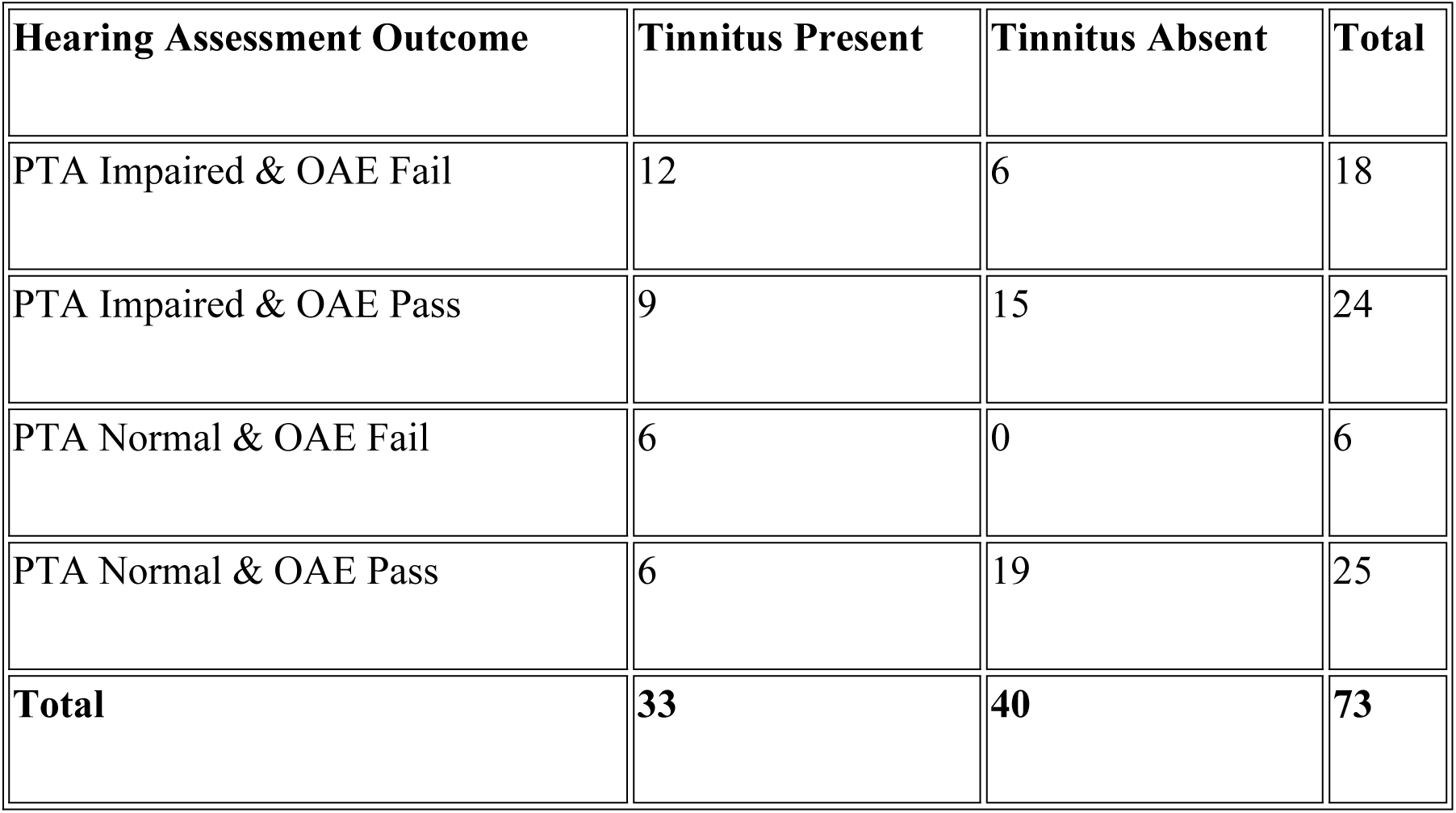
Cross-Tabulation of Pure-Tone Audiometry (PTA) and Distortion Product Otoacoustic Emission (DPOAE) Results by Tinnitus Status among Generator Technicians.

| Hearing Assessment Outcome | Tinnitus Present | Tinnitus Absent | Total |
| --- | --- | --- | --- |
| PTA Impaired & OAE Fail | 12 | 6 | 18 |
| PTA Impaired & OAE Pass | 9 | 15 | 24 |
| PTA Normal & OAE Fail | 6 | 0 | 6 |
| PTA Normal & OAE Pass | 6 | 19 | 25 |
| **Total** | **33** | **40** | **73** |

## DISCUSSION

This community-based pilot study revealed a prevalence of tinnitus (45.2%) among informal generator technicians in metropolitan Northern Nigeria, an occupational cohort often neglected in hearing health research and policy discussions. This finding underscores the urgent need to reframe tinnitus not just as an auditory complaint, but as a sentinel symptom of deeper population-level vulnerabilities related to unregulated noise exposure.

The strong association between prolonged occupational duration (≥10 years) and reported tinnitus points to the cumulative risk posed by chronic exposure to high-decibel generator noise. Such exposure, routinely exceeding 90 dB(A), has been documented in informal trades across Nigeria and sub-Saharan Africa.^[17, 19]^ These workers typically lack access to protective equipment, routine hearing screening, and noise hazard education, which places them in what may be described as “high-risk low-protection zones” within urban economies. Our findings support calls for formal recognition of informal workers in national occupational safety frameworks, especially given the scale and importance of the generator repair sector in powering local commerce or small and medium scale enterprises.

Notably, OAE test failure emerged as a significant independent correlate of tinnitus. Table 4 further illustrate that tinnitus can occur with normal audiometric thresholds but failed OAE results. OAE appears to offer greater sensitivity than conventional pure-tone audiometry in detecting early-stage auditory dysfunction, even in those without overt hearing loss.^[25, 26]^ This is clinically relevant as it suggests that OAE screening could serve as a cost-effective, field-friendly tool for identifying subclinical cochlear damage in settings where access to audiological infrastructure is limited or absent. Integrating portable OAE devices into community health outreach or primary care screening initiatives may help bridge this diagnostic gap for underserved noise-exposed populations.

Interestingly, audiometric impairment did not retain significance in multivariable analysis. This may reflect the well-documented phenomenon where tinnitus can occur independently of hearing threshold shifts, suggesting a disconnect between subjective auditory perception and measurable hearing loss. Several studies have also emphasized the role of central auditory plasticity and neural hyperactivity in the generation of tinnitus, even in individuals with clinically normal audiograms.^[7, 27, 28]^

Though age was not significantly associated with tinnitus in the adjusted models, the distribution in table 3 reveals symptom presence even among adolescents and younger adults. This underscores the need for early occupational health education targeting youth in informal trades. The initial association of marital status with tinnitus raises questions about psychosocial mediators. While it lost significance after adjustment, it’s plausible that married individuals, particularly in cultural contexts emphasizing familial interdependence, are usually more conscious of bodily symptoms and/or encouraged by spouses to seek care.

Emotional awareness, stress exposure, and social support structures may influence the likelihood of symptom reporting, a trend observed in tinnitus studies conducted in both Western and African populations.^[29, 30]^ These dimensions need further investigation using mixed-method approaches to capture lived experience and behavioural drivers of auditory symptomatology.

From a public health perspective, our study’s findings hold broader implications. Tinnitus is increasingly linked with mental health disorders, including sleep disturbance, anxiety, and depression.^[10–12, 31]^ Informal sector workers, many of whom face economic challenges, occupational instability, and environmental hazards, may be at heightened risk for psychological distress exacerbated by persistent tinnitus. This also calls for integrated care models in primary health systems, where auditory screening is coupled with mental health triage in resource-constrained settings.

Another important yet under-discussed finding is the presence of participants as young as 14 years. Early entry into generator-repair work exposes adolescents to harmful acoustic environments at a critical developmental stage. This raises ethical and regulatory questions about child labour and occupational health education for youth in informal trades. Preventive health campaigns tailored to younger workers could deliver long-term dividends in reducing lifetime auditory impairment.

The economic ripple effects of hearing loss, including reduced productivity, diminished communication efficiency, and increased healthcare costs, are well-documented.^[32]^ In skill-based vocations like generator repair, auditory acuity is not just biologically significant, it is economically essential. Thus, untreated tinnitus and progressive hearing loss could threaten the livelihood of thousands of informal workers.

Despite these insights, this study has several limitations. The sample size, while adequate for a pilot, limits generalizability and statistical power for subgroup analysis. The cross-sectional design precludes causal inference and cannot track progression of tinnitus over time. Tinnitus assessment was based on self-report without the use of validated instruments, which may introduce measurement bias. Only male participants were enrolled, reflecting occupational demographics but limiting gender-based extrapolations. Additionally, confounding factors such as medication history, mental health status, smoking, and recreational noise exposure were not controlled for. Finally, although OAE testing was conducted using a portable screener in a sound-treated room, this equipment may offer reduced sensitivity compared to full clinical OAE systems.

Finally, this study highlights an urgent public health need: recognizing and addressing tinnitus as a proxy for occupational auditory risk in Nigeria’s informal economy sector. With nearly half of generator technicians reporting symptoms, and subclinical cochlear damage detectable via OAE testing, this pilot points toward scalable interventions, ranging from mobile hearing screening units to hearing protective device advocacy and occupational noise regulation.

## CONCLUSION

This study found that tinnitus affects nearly half of informal generator technicians in urban Kano-Nigeria, with prolonged occupational noise exposure and OAE failure serving as significant predictors of the condition. These findings highlight both the prevalence of cochlear dysfunction and the potential of OAE screening to detect auditory damage earlier than conventional methods. Given the limited health safeguards in informal work environments, integrating hearing protection, noise education, and portable OAE testing into community outreach programs presents a practical and culturally responsive strategy.

Expanding auditory health surveillance, especially for youth entering noisy trades e.g. generator repair settings, could help preserve long-term functional capacity, economic productivity, and social wellbeing. Future longitudinal studies would examine symptom trajectories, broader systemic barriers to auditory health equity across informal labour populations, tinnitus progression, gender differences, and the impact of targeted interventions tailored to informal sector dynamics in low-resource settings.

## Acknowledgements

The authors extend their sincere gratitude to the leadership and members of the Generator Mechanics Union in Kano for their invaluable collaboration throughout the study.

Appreciation is also due to the audiology team at the ENT Clinic, Aminu Kano Teaching Hospital, whose technical expertise and support during data collection were instrumental to the success of this research.

## Author information

### Contributions

FIT-A conceived the study, FIT-A, NB-M and AA contributed to the interpretation of data, revision of the article critically for important intellectual content, and approved the final version of the manuscript. AA also supervised the study. The corresponding author, attest that all listed authors meet authorship criteria and that no one meeting authorship criteria was omitted.

### Authors’ information

The authors are entirely responsible for the opinions or views expressed in this article and do not represent the views, decisions, or policies of their various affiliated institutions or any government body.

#### Consent for publication

Not applicable

### Ethics approval and consent to participate

Ethical approval for this study was obtained from the Kano State Ministry of Health Ethics Committee (NHREC/17/03//2018) and the Bayero University Health Research Ethics Committee (NHREC/BUK-NHREC/348/10/2311). In addition, permission and support to conduct the research were secured from the leadership of the Generator Mechanics Union. Written informed consent was obtained from all individual participants prior to enrolment. The data analysed are from the Occupational Noise Induced Hearing Loss project, Generator mechanics data. The study was conducted in accordance with the ethical principles outlined in the Declaration of Helsinki, as revised in 2013.

### Data Availability

The datasets generated and analysed during the current study are available from the corresponding author on request.

